# A Qualitative Exploration of the use Social Media for Learning by Emergency Medicine Doctors

**DOI:** 10.1101/2024.11.17.24317333

**Authors:** Sarah Edwards, Nabilah Mayat

## Abstract

**Background:** The use of social media for learning amongst emergency medicine doctors has been increasing in recent years. However, the role of social media as a learning modality in emergency medicine is not fully understood. This study aims to understand why and how emergency medicine doctors use social media for learning. Through this understanding this study aims to promote thought into design of future resources of learning on social media.

**Methods:** Emergency Medicine doctors of all grades were recruited, from a single tertiary University Hospital in the United Kingdom. These participants were interviewed by semi structured interview following the principles of a Critical Incident Technique. Recordings were professionally transcribed. Transcriptions were then thematically analysed and coded to understand participant’s perspectives. Ethics was granted by Cardiff University, Wales.

**Results:** 20 emergency medicine doctors from Foundation Year 1 through to consultants were interviewed. Twitter (X) was the most popular platform; infographics were the most popular resource and 80% spent less than 3 hours per week on SM for learning. Key themes from the thematic analysis as to why emergency medicine doctors used social media for their learning included wanting to keep up to date ease of access, discussion around topics, bench marking current practice, community, and recommendations. Only 10% used social media as part of their continued professional development and 65% stated social media had influenced their clinical practice.

**Conclusions:** This unique qualitative explorative piece of work explores the use of social media for learning by emergency medicine doctors and how it is changing clinical practice.

With the rapid growth and uptake in social media use for educational purposes. It is vital that all educators fully understand how it is being currently used as well as views of learners on this form of learning. This will allow us to further grow and develop this exciting area and maximise the educational impact on its users.

**What is already known about the subject?:**

- The use of social media for learning is popular amongst emergency medicine doctors
- The term FOAMed was first created following an Emergency Medicine Conference in 2012.
- There are increasing amounts of FOAMed resources produced each year related to emergency medicine.

**What this study adds?:**

- X (Twitter) is a popular social media platform used by doctors
- Information learnt from social media is being applied to clinical practice
- Social media is being used for many reasons including to keep up to date, discussion around topics and bench marking of current practice with others
- Learning through social media is being increasingly recognised as a form of Continued Professional Development (CPD) yet commonly not reported by learners as such.

## Introduction

Medical Education is a constantly evolving field. A world where the treadmill of the creation of new learning resources and innovations never ceases. The current generation of Medical Education often experiences resources relayed in digital formats through the medium of social media (SM). ^1,2^As the world of SM has developed, these communications are seen to occur across “platforms,” examples of which include Facebook, X (Twitter), Instagram and Reddit.

The ‘Free Open Access Medical Education’ (FOAM) movement is an example of a pathway through which these resources can be disseminated amongst healthcare professionals. This relay of resources occurs using an identifier tagged to each resource, specifically, the hashtag #FOAMed. This identifier enables localisation and therefore, collation and easy access to an increasing pool of learning resources on social media. Whilst its concept has been around for many years, the term ‘FOAM’ was coined by Mike Cadogan via the Life in the Fast Lane Blog and first discussed in the international conference of Emergency Medicine (ICEM) 2012 in Dublin^3^. FOAM resources created and shared on SM in the name of medical education could be classed as social media Medical Education (SoME) resources.

Within the vast amount of available ‘FOAM’ resources, there is an ever-growing pool of Emergency Medicine resources.^4^ In busy specialities with ever increasing pressure on medical professionals such as Emergency medicine (EM), keeping up to date can be an incredible challenge especially due to the breadth of knowledge needed to practice. Following the COVID-19 Global Pandemic in early 2020 saw a global shift of medical education shift to online, especially increasing use of SM for learning.^5–8^ Subsequently, as the world began to open again the role of SM in learning is yet to be determined.

However, what does continue is the concept of #FOAMed. With the role of SM has had in recent years it is important to appreciate how it is clear to see that this world of SM has a clear role in the world of emergency medicine education and learning in Medical Education in general. However, how well do we understand this role and the world of learning on SM in general? Are we truly aware of if people are learning on SM and if so, what, and how are they learning?

From anecdotal experiences, we, the authors have seen a rise in EM practitioners utilising social media. However, we note a lack in literature further exploring this phenomenon limiting our understanding of key factors such as active and passive learning on SM and appealing factors of this form of learning amongst EM doctors. This study aims to explore this phenomenon to better understand the world of SM in medical education especially amongst EM practitioners. Through understanding social media use in EM learners, we aim to explore how this newfound understanding can help enhance learning on social media both for EM practitioners as well as beyond the world of EM. We believe it is only through building this understanding that learning on SM can be optimised especially as this form of learning is only rising with time.

## Methodology

### Aims

This qualitative study aimed to develop understanding of participants’ perspectives and experiences of using SM for learning in medical education. Through this understanding, this study aims to facilitate exploration of how best SM based learning can be optimised with consideration of other specialities as well as EM. This will be explored through understanding the following:

- Key factors driving learning on social media
- Appealing aspects of preferred learning resources on social media
- Effects of learning on SM as perceived by the learner

### Setting

This study was performed at Leicester Royal Infirmary’s Emergency Department, a large university teaching hospital in the United Kingdom with 2000 beds, employing 150 medical staff with a catchment population of 1.1 million. It is a hospital operating across 3 sites consisting of collocated emergency medicine ie. Not a Level 1 Trauma Centre.

### Research Paradigm

This qualitative research is based on constructivist epistemology; exploring what it assumes to be a socially constructed dynamic reality^9^. This occurs through a flexible, descriptive, holistic and context sensitive way. Therefore, it is an in-depth description of what the phenomenon being researched is, from the perspectives of the people involved^9^. It tries to understand the social experience, giving it meaning whilst appreciating that reality or knowledge are socially and psychologically constructed. Therefore, the researcher is supposed to develop a close relationship with the subjects being studied^9^.

To understand this human behaviour in depth, this work will focus on participant lived experience through an ethnographic lens by aiming to “get inside” and understand their thoughts^9^. This aim was made more possible through the fact that the researcher, who also acted as interviewer, SE, was a colleague of participants interviewed and therefore, more likely to be able to access that insider knowledge.^10^

### Participants and recruitment

The recruitment process for this study aimed to recruit ten EM doctors at various grades. Eligibility Criteria for participation were for participants to be EM or PEM Clinicians. The topic guide and interview schedule were piloted with two participants. This pilot worked to demonstrate the topic guide and interview schedule as fit for use to answer the aims of this study. Participants were initially recruited using convenience sampling through internal staff email advertisements. Participants gave consent for recruitment. No patients or public were recruited as part of this study.

The sample size was selected based on anticipated feasibility of recruitment, representing approximately ten percent of the clinical workforce as the recommended pilot group size by Guest et al. (2006)

### Data collection

All interviews were conducted by one researcher, Sarah Edwards (SE). Interviews were semi-structured using a topic guide and were audio recorded. Recordings were professionally transcribed using Rev.com^11^. Interviews took place face-to-face, with only SE, in non-clinical environments in the hospital and lasted up to thirty minutes. There were no repeated interviews, notes were taken if needed during the interview. Participants were not given copies of their interviews.

The semi structured interview followed the principles of a Critical Incident Technique (CIT)^12^. CIT allows researchers to examine behaviours to understand practices and system changes that may need to occur because of it^12^. The semi structured interviews were shaped to allow for CIT, wherein a ‘critical incident’ is defined as an action that contributes to an effective outcome or the inverse.^12^ It is the recognition of such ‘critical’ incidents when talking to participants that facilitated shaping the semi structured interviews through the ‘people’s perspectives’ in line with the constructivist epistemology of this study.

### Data Analysis

Interview transcripts were read and re-read by SE to check for transcription accuracy and to improve familiarisation with the data. Transcription data was organised in NVivo (version 11)^13^. The data was open coded using an inductive approach. Themes were not planned. Codes reflected quotation themes, for instance ‘learning from newspapers’ and ‘reflecting on other, clinicians care’. An inductive coding approach was undertaken. This is where codes are generated from the data, in a ground up approach. Codes were condensed and merged to form themes for each research question, which were constantly compared and iteratively edited as new data codes were added. Participants were not asked to comment on the results.

### Ethics and funding

Ethical approval was granted by Cardiff University on 6^th^ February 2019 (reference 19/02). The researchers received no specific funding for this study.

### Reflexivity and Bias

SE is a female interviewer employed as an education fellow in the department at which this study was held at the time of the study. Peer interviewing has been shown to produce greater depth of insight due to being an “Insider” and thus, a trusted researcher and thus more beneficial from an ethnographic perspective. However, peer interviewing can also add unexpected bias to projects.^14^ Therefore, care was taken to maintain a reflexive approach through discussions with supervisors and self-reflection. Participants were reminded that this study was anonymous, confidential and would have no bearing on their work within the department.

## Results

### Number of participants

21 doctors spanning grades from recently qualified to consultant (See Table 3 or Table 1.1 in Supplementary Material), were interviewed in February 2019.

Given this was a study with the use of interviews generating qualitative data to be analysed, the sample size of 20 people was deemed to achieve data saturation. Data saturation is defined as consistent repetition of expressed ideas and the absence of new phenomena and was sought among medical education themes.^15^

Of the 21 Participants, 20 Interviews were included in data analysis. No participants dropped out. One Participant was excluded (P8) as it became apparent that the Participant used SM for a focus of other than for their own learning (Use of SM to educate).

### Participant demographics and time spent on social media

Table 1.1 (Supplementary) shows participant demographics including grade, age range and amount of time spent on SM for learning per week. Across all the participants, representation of similar numbers across all grades were found. Seventy percent (70%) of participants were 35 years of age or less. Eighty percent (80%) of the participants spent 3 hours or less per week on SM for learning.

### Thematic Analysis

A thematic analysis was carried out to ascertain the key driving factors for learning, illustrated in Table 2 below followed by a further exploration of these themes.

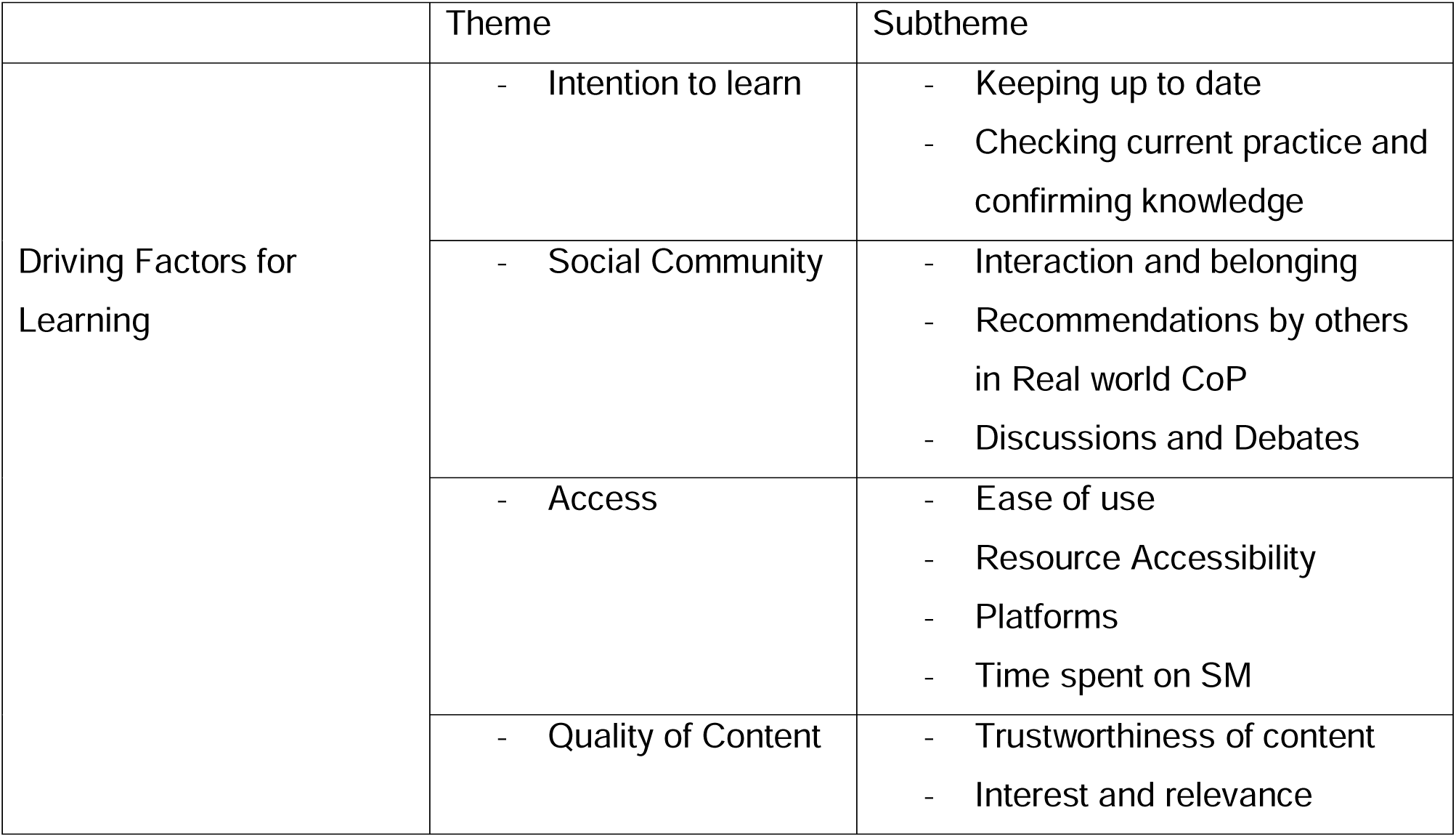
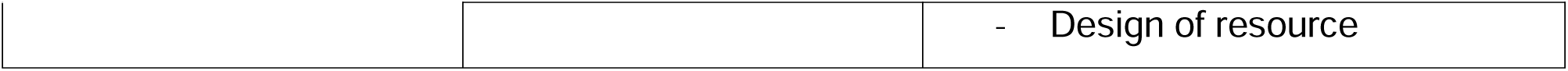

|  | Theme | Subtheme |
| --- | --- | --- |
| Driving Factors for Learning | - Intention to learn | - Keeping up to date<br>- Checking current practice and confirming knowledge |
|  | - Social Community | - Interaction and belonging<br>- Recommendations by others in Real world CoP<br>- Discussions and Debates |
|  | - Access | - Ease of use<br>- Resource Accessibility<br>- Platforms<br>- Time spent on SM |
|  | - Quality of Content | - Trustworthiness of content<br>- Interest and relevance |
|  |  | - Design of resource |

### Themes (Table 3)

| Subtheme | No. of Participants | Roles |
| --- | --- | --- |
| - Keeping up to date | 7 (35%) | All except Consultants |
| - Checking current practice and confirming knowledge | 6 (30%) | Majority senior grades |
| - Interaction and belonging | 5 | All grades |
| - Recommendations by others in Real world CoP | 6 | ---- |
| - Discussions, Questions and Debates | 4 | Early career upto ST4 |
| - Ease of use | - | - |
| - Resource Accessibility | - 16 (80%) | - All grades |
| - Platforms | - | - |
| - Time spent on SM | - | - |
| - Trustworthiness of content | - 7 (35%) | - Mostly Junior Grades |
| - Interest and relevance | - 10(50%) | - All grades |
| - Design of resource | - 8 (40%) | - |

### Theme 1 – Intention to Learn

Participants approached SM with the intention to learn through expanding horizons with new knowledge as well as gaining reassurance through checking and alignment of practice and knowledge with peers.

> *“I definitely have found that I became more current when I was using Twitter™ and got a greater breadth of what was going on, and development both UK and abroad” ST5 EM – P1*

> *“So, there’s stuff that is completely new to me, but what I like to know is that my peers or colleagues from around the world are doing similar things.” Consultant in Paediatric EM – P13*

Of the 20 participants involved in this work, 13 (65%) could identify practices that they had adopted from SM in their clinical practice. A key example, mentioned by 4 participants and the most referenced resource amongst participants, was the modified Valsalva manoeuvre from the REVERT trial, used in the management of supraventricular tachycardias^16^.

> *“Probably the best one would be something like the modified valsalva from the REVERT Trial, where actually they’re tweeting about a significant difference, and because of that significant difference you’re more interested to go and find the primary literature…. this I adopted into my clinical practice” ST5 EM – P1*

Whilst all these participants clearly use SM for their learning, only 2 participants (10%) recorded this formally down as part of their Continued Professional Development (CPD). Key reasons behind this included uncertainty surrounding eligibility of SM based learning as. CPD and lack of time to officially document learning.

### Theme 2: Social Community

Community introduced learners to SM with participants exploring learning on SM through recommendations from colleagues in their real-world Community of Practice to deciding to continue learning on SM due to the virtual communities and communal spirit established through such platforms.

> *“It’s more of a communal learning sort of thing”. ST6 EM – P19*

The ability to have discussions and questions answered through this community was of particular appeal.

> *“I like the fact that there’s a bit of a discussion around what the learning is about, and it’s a dynamic thing as opposed to just, I don’t know, reading something and then that’s it. You read something and then there’s a discussion about it, and then people add points into the learning, which you might not have otherwise had.” ST3 EM*

### Theme 3: Access

Access to resources influenced learning on SM with participants valuing resources which were free, convenient, and easily accessible. Twitter™ was by far the most popular SM platform, accessed by 75% of participants followed by Facebook™ and YouTube™. 

> *“Textbooks are heavy. I can’t be bothered to move them around… It’s much easier to read something that is online” Consultant in Paediatric EM – P13*

> *“It’s available, updated. It can be used anywhere I am, always on my telephone, which is really very convenient, mobile and everywhere, every time” Non-training CT1 Level*

### Theme 4: Quality of Content

Participants assessed the quality of content they used for learning on SM through evaluating how interesting and trustworthy they perceived resources to be. This evaluation of perceived trustworthiness considered visual and knowledge-based aspects.

> *“The people who are doing that are really trusted people, scientific, organised, and updated” ST1 Level Doctor – P7*

Visually appealing resources with a good foundation of knowledge were valued amongst 40% of participants with resources displaying good summaries of a topic or an interesting topic being deemed as knowledgeable. Participants interacted with varying forms of content including podcasts, blogs, websites, and infographics. Of these, infographics were the most popular, used by 11 Participants. 20% of participants found summaries to enhance learning.

> *“Okay. So, let’s be honest, a picture with a tweet instantly makes it more interesting, purely because it stands out of a news feed that’s otherwise mostly text based” ST5 EM*

However, not all participants adhered to such a targeted approach with seven participants of all grades except consultants choosing to learn through a process of random resource selection. Factors influencing some participants to be more targeted as opposed to random in their approach were unclear.

> *“To be honest, I don’t really make a conscious decision as to what I use.” ST3 EM – P6*

## Discussion

In 2012 the average person spent 10.5 hours per week using SM, this had increased to 17.6 hours per week in 2023.^17^ The results of our study showed SM use amongst EM doctors to be at a similar level. This study set out to explore SM use by EM doctors for learning. 80% of participants used SM for up to 3 hours of learning online with the findings of Mallin (2014), who found at least 97.7% of participants spent at least 1 hour per week using SM for learning^18^.

Through this piece of work, four key themes were found to drive learning on social media (See table 2); Intention to Learn, Social Community, Quality of Content and Access. To the best of our knowledge, this piece of work is unique in demonstrating the real-world effects of learning virtually on social media, with participants demonstrating a change in clinical practice.

Despite the real-world impact on clinical practice, this piece of work also interestingly found only 2 participants to formally reference this form of learning as CPD. One of the recurring issues around the use of SM resources for CPD, is around the perceived reliability and credibility of it. The General Medical Council (GMC) is the national licensing body for doctors in the UK. Doctors in the UK are required to engage in CPD^19^similarly to other doctors globally. CPD as defined by the GMC is “any learning outside of undergraduate education or postgraduate training that helps you maintain and improve your performance”.^19^ Participants noted being unaware of learning via SM as a valid form of CPD. This is despite the Royal College of Emergency Medicine (RCEM) in the UK allowing the use of SM learning to be used as evidence for CPD^20^. It would be interesting to consider whether this lack of inclusion of SM driven learning as part of CPD is due to the inherent perception of trainees of the credibility of learning via SM or whether it is due to a lack of awareness that SM is now recognised as a legitimate source of CPD learning.

65% of participants reported SM to influence their clinical practice. Given that we recognise the importance of SM as a tool which can be used for both good and bad, it would be useful to consider the effect of this learning on clinical practice. Given SM is increasingly being used to disseminate publications, it is plausible that this results in a faster integration of latest research informed practice from a change over decades to one that occurs over years. This knowledge translation from SM is ever present being used extensively by the whole spectrum of undergraduates through to postgraduate doctors.^21^ However with information spreading fast, such as in the COVID 19 pandemic, it is ever important to understand the credibility behind information presented as ‘latest research’.^22,23^

Participants were found to evaluate the resources they chose to use on SM for learning. Key valued aspects consisted of visual appeal and perceived trustworthiness of resources. Infographics were the most common resource that people interacted with on SM within this work, followed jointly by podcasts and blogs. The concept of an infographic has been around for 20000 years, with the mainstream word becoming more popular in 2010^24^. Whilst infographics are not mentioned in the EM literature as a resource used, blogs and podcasts are cited as popular learning resources that EM doctors use^4,25–28^. It is unclear as to why this difference is present amongst resources.

Resources for learning were also identified through the surrounding context, both through platforms as well as communities of individuals enabling learning through discussion. X (Twitter) was by far the most popular SM platform used for learning with 75% of interviewed participants utilising it. This contrasts with what other studies from the EM literature have found^4,27,28^. Using surveys to understand SM platforms used by EM doctors, Pearson (2015) found Facebook followed by Youtube to be the most popular. Purdy (2015) and Folkl (2016) found differing results with Youtube™ and GooglePlus most used. In contrast, literature investigating SM use specifically for learning showed X (Twitter) as the most popular platform amongst doctors interviewed. ^29–31^ When considering these differences in findings, we hypothesise this as evidence to the ever-evolving nature of the world of SM. We feel these differences in platform usage reflect evolution of SM with time especially due to the differing years of studies. However, we cannot definitively attribute evolution of SM to account for these differences in findings. With Twitter having renamed itself as X, and a sense that the world of #MedTwitter is in turmoil.^32^ How this reinvention of the platform will affect learning, is yet to be determined.

To the best of our knowledge, this is the first interview-based study exploring SM for learning amongst EM doctors. It is our understanding that the qualitative nature of this study is what allowed us to understand key themes in depth. Furthermore, the use of “insider research” to obtain an ethnographic element to the research, has enabled increased depth of insight into participant experience.^10^

### Limitations

Given that this work only relates to EM and PEM doctors, it would be difficult to predict how transferable the findings of this piece of work are for doctors in other specialities. However, doctors in General practice, Acute general medicine, Acute general paediatrics, and those at the beginning of their training, all would generally find themselves in similar contexts to those in EM and thus, it is possible that findings could be transferable to these individuals in a broader sense.

This study was carried out with all participants practicing in one hospital in the UK. This could further limit the transferability of findings from this study. However, due to the data rich methods used along with a methodology focused on individual lived experience, we envisage the data achieved through this study to still be of use in furthering our knowledge of this phenomenon. Although lived experience cannot be completely reproduced, to confirm validity of the results and general applicability interviewing other doctors with different lived experiences from other centres in the UK would be a useful comparison. When considering these findings more globally interviews with doctors from other countries would give even further insight especially given the global nature of SM.

CIT is a flexible approach that can be adapted to each situation. The semi structured approach allowed for consistency of information collected, whilst the CIT method ensured as much information to understand the participant’s behaviours. The analysis of the interviews sought to retain the close connection between the participant’s comments within the context of the whole interview. However, there are limitations to this technique that includes bias to the most recent events happening. Also, inherently this technique relies on people’s memories to recall this event and accuracy can be affected by this. Despite this, the large data set may have offset some of these limitations

This piece of work aimed to use ‘insider research’ to bring a new ethnographic perspective to researching the lived experience of participants. Although the use of insider research can result in collection of more in-depth unique data, it would also be important to acknowledge the possibility of confirmation bias and of peer interviewing affecting results.^33,34^

There has been a delay of this work being published due to the COVID pandemic. However, the unique content contained within this work has not ultimately changed. With COVID decreasing possibility of group based and face to face learning, learning from SM has increased^23^. We have progressed from a world of predominantly face to face teaching to virtual during the pandemic, to now progressing to essentially a new style of medical education, a hybrid between the worlds of virtual and in person learning. As stated by Merchant and Lurie (2020) and Katz and Nandi (2021)^18^, the use of social media in allowing physicians to share learning and conversations highlights the importance of how physician engage with learning and information on social media. This work can add into this bank of knowledge and highlights the increasing importance in understanding this form of learning.

## Conclusion

Emergency medicine doctors have been found to be using social media to support their learning which has in turn influenced clinical practice for many of these physicians. Therefore, this study highlights the importance of and aims to understand the lived experience of learning on social media. Key themes as to why emergency medicine doctors used social media for their learning included wanting to keep up to date, ease of access, discussion around topics, benchmarking current practice, community, and recommendation.

As we move into an ever-evolving virtual world impacted lately by the post Covid pandemic stage of learning, this study may help resource development and delivery of future learning for doctors on social media. It is our hope that this understanding can help achieve a world where SM is a beneficial tool for learning and therefore, creating growth in clinical practice.

## Data Availability

All data produced in the present study are available upon reasonable request to the authors

## Appendix 1 Questions used in Interviews

### Questions for Semi Structured Interview – MSc Medical Education: Exploration of the use of social media by emergency medicine doctors for learning; a small-scale study

Thank you very much for your time. You are free to stop this interview at any time and you are free to withdraw from the study at any time. I am going to ask you some questions and record the interview.

During this interview we will be discussing your use of social media. When referring to social media I will be referring to the platforms such as Facebook, Twitter, Instagram, Reddit. Resources from these platforms include blogs, infographics, threads, podcasts, online discussions, posters.

General Information

1. What grade are you?
2. Are you an emergency medicine trainee? or is this a rotational post?
3. If applicable: how long have you been working in emergency medicine (EM)?

General information about Social Media

1. Do you use Social Media to facilitate your professional development?
2. Can you tell me what social media platforms you use for learning?
3. Can you tell me about your use of Social media for your learning?
4. What type of social media resources do you use for your learning? Blogs, infographics, podcasts etc.
5. How do you decide what resources you use?
6. Why do you use those resources?

Influence of Social Media on Clinical Practice and CPD

1. Can you remember the last social media education resource you used/ interacted with?
2. What type of resource was this?
3. Can you relive the decision-making process as to why you interacted with this resource?
4. Did that resource influence your clinical practice?
5. Have there been any other examples of SoME influencing your clinical practice and you tell me about that please?
6. How did the SoME post change your clinical practice?
7. Do you use SoME as part of you CPD?

Participant Demographics:

**Table 1.1:**
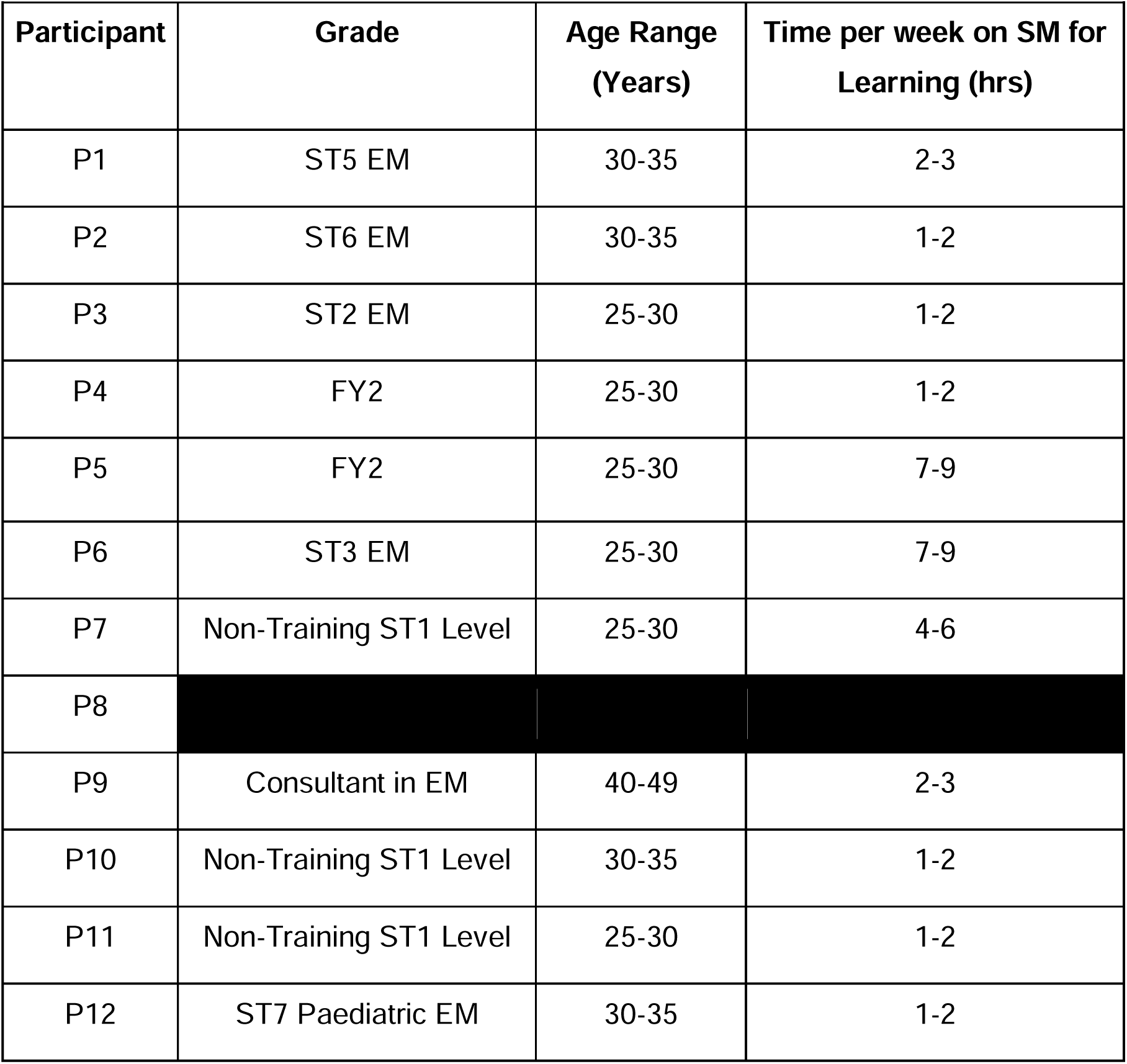
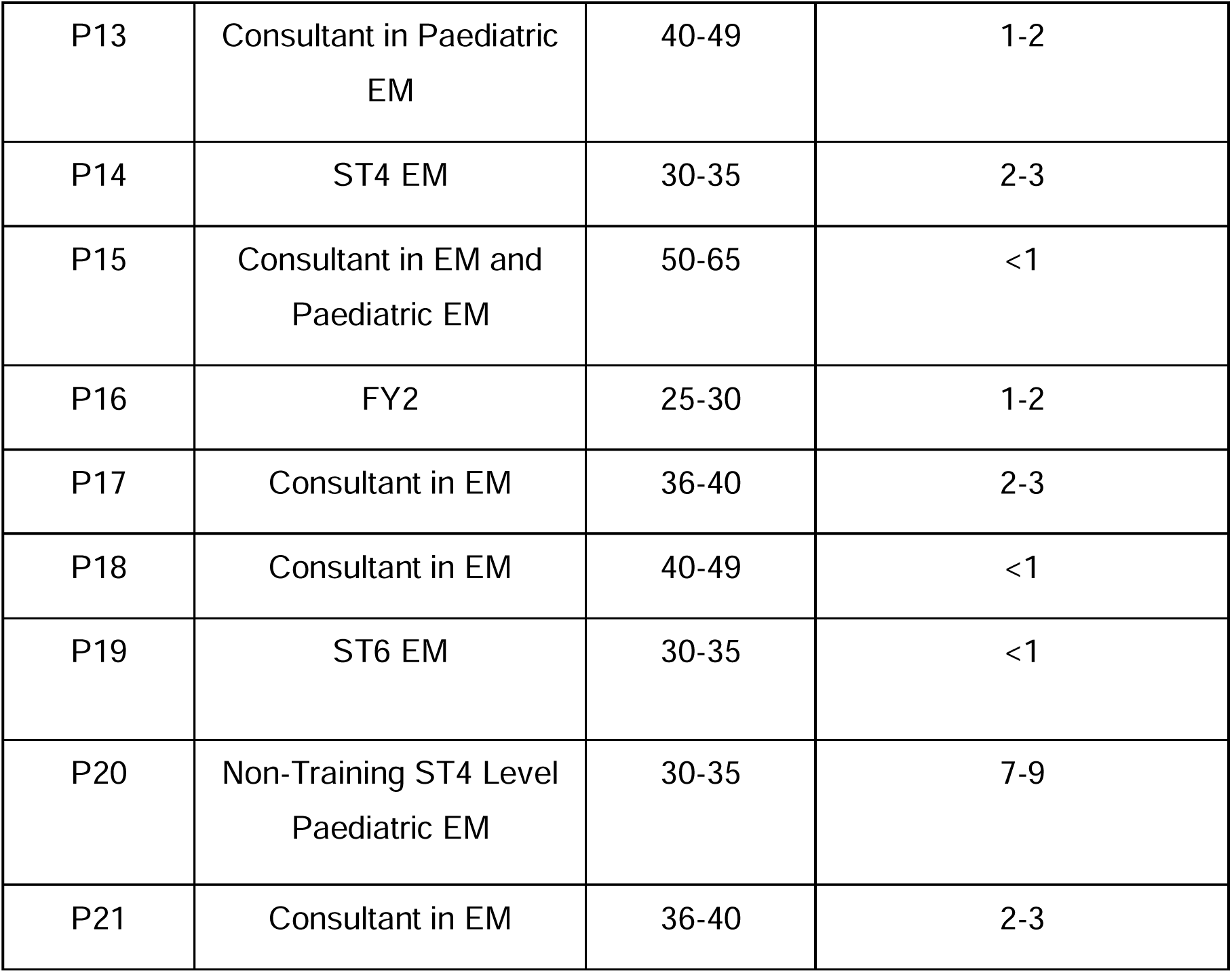
Participant demographics and time spent on social media.

| **Table 1.1: Participant demographics and time spent on social media** |  |  |  |
| --- | --- | --- | --- |
| **Participant** | **Grade** | **Age Range (Years)** | **Time per week on SM for Learning (hrs)** |
| P1 | ST5 EM | 30-35 | 2-3 |
| P2 | ST6 EM | 30-35 | 1-2 |
| P3 | ST2 EM | 25-30 | 1-2 |
| P4 | FY2 | 25-30 | 1-2 |
| P5 | FY2 | 25-30 | 7-9 |
| P6 | ST3 EM | 25-30 | 7-9 |
| P7 | Non-Training ST1 Level | 25-30 | 4-6 |
| P8 |  |  |  |
| P9 | Consultant in EM | 40-49 | 2-3 |
| P10 | Non-Training ST1 Level | 30-35 | 1-2 |
| P11 | Non-Training ST1 Level | 25-30 | 1-2 |
| P12 | ST7 Paediatric EM | 30-35 | 1-2 |
| P13 | Consultant in Paediatric EM | 40-49 | 1-2 |
| P14 | ST4 EM | 30-35 | 2-3 |
| P15 | Consultant in EM and Paediatric EM | 50-65 | <1 |
| P16 | FY2 | 25-30 | 1-2 |
| P17 | Consultant in EM | 36-40 | 2-3 |
| P18 | Consultant in EM | 40-49 | <1 |
| P19 | ST6 EM | 30-35 | <1 |
| P20 | Non-Training ST4 Level Paediatric EM | 30-35 | 7-9 |
| P21 | Consultant in EM | 36-40 | 2-3 |

